# An investigation of age-related neuropathophysiology in autism spectrum disorder using fixel-based analysis of corpus callosum white matter micro- and macrostructure

**DOI:** 10.1101/2022.03.30.22273215

**Authors:** Melissa Kirkovski, Mervyn Singh, Thijs Dhollander, Ian Fuelscher, Christian Hyde, Natalia Albein-Urios, Peter H Donaldson, Peter G Enticott

**Author notes:** Corresponding author Dr Melissa Kirkovski.

## Abstract

**Background:** Corpus callosum anomalies are commonly noted in autism spectrum disorder (ASD). Given the complexity of its microstructural architecture, with crossing fibers projecting throughout, we applied fixel-based analysis to probe white matter micro- and macrostructure within this region. As ASD is a neurodevelopmental condition with noted abnormalities in brain growth, age was also investigated.

**Methods:** Data for participants with (N=54) and without (N=50) ASD, aged 5-34 years, were obtained from the Autism Brain Imaging Data Exchange-II (ABIDE-II). Within each site, indices of fiber density (FD), fiber cross-section (FC), and combined fiber density and cross-section (FDC) were compared between those groups.

**Results:** Young adolescents with ASD (age = 11.19 ± 7.54) showed reduced macroscopic FC and FDC compared to age-matched neurotypical controls (age = 10.04 ± 4.40). Reduced FD and FDC was noted in a marginally older ASD (age 13.87 ± 3.15) cohort compared to matched controls (age = 13.85 ± 2.90). Among the oldest cohorts, a non-significant trend indicated reduced FD in older adolescents/young adults with ASD (age = 17.07 ± 3.56) compared to controls (age = 16.55 ± 2.95). There was a positive correlation between age and callosal mean FC and FDC in the youngest cohort. When stratified by diagnosis, this finding remained only for the ASD sample.

**Conclusion:** White matter aberration appears greatest among younger ASD cohorts. In older adolescents and young adults, less of the corpus callosum seems affected. This supports the suggestion that some early neuropathophysiological indicators in ASD may dissipate with age.

## Introduction

Autism spectrum disorder (ASD) is an etiologically complex, pervasive, and life-long neurodevelopmental condition characterized by social impairments and the presence of restricted and repetitive patterns of behavior and interests (RRBI) (American Psychiatric Association, 2013). There is strong evidence for structural and functional neuropathophysiology underlying ASD symptomatology (Girault & Piven, 2020; Philip et al., 2012; Sato & Uono, 2019; Wass, 2011).

The corpus callosum (CC) is the largest commissural tract in the brain and is one of the most commonly implicated white matter (WM) tracts in research investigating the structural neuropathophysiology of ASD. The CC has a crucial role in interhemispheric signal transfer, or connectivity, and tracts between the frontal, parietal, temporal, and occipital lobes, both cortically and subcortically, project through the CC (De Lacoste, Kirkpatrick, & Ross, 1985; Hofer & Frahm, 2006; van der Knaap & van der Ham, 2011). Importantly, aberrant interhemispheric connectivity is commonly reported in ASD populations (Anderson et al., 2011; Dickinson et al., 2018; Guo et al., 2020; Li et al., 2019; Zhu, Fan, Guo, Huang, & He, 2014). Morphologically, it is generally established that CC volume and thickness are reduced in ASD groups compared to matched controls (Anderson et al., 2011; Frazier, Keshavan, Minshew, & Hardan, 2012; Freitag et al., 2009; Hardan et al., 2009; Li et al., 2019; Temur et al., 2019), though the evidence is mixed (Kucharsky Hiess et al., 2015; Lefebvre, Beggiato, Bourgeron, & Toro, 2015). Diffusion MRI (dMRI) allows for a deeper understanding of the WM micro- and macrostructural effects that might contribute to these reductions in ASD. Early dMRI based research generally indicates that fractional anisotropy (FA) is reduced in ASD compared to controls. Mean-(MD), axial-(AD), and radial-diffusivity (RD), in contrast, are generally reported to be increased at the CC of individuals with ASD (Yao, Becker, & Kendrick, 2021). There is, however, a great deal of variability in the literature. Numerous studies have reporting opposing findings to those described above, and many report no differences in WM microstructure in ASD compared to neurotypical (NT) cohorts (for comprehensive reviews refer to: Aoki, Abe, Nippashi, & Yamasue, 2013; Hoppenbrouwers, Vandermosten, & Boets, 2014; Travers et al., 2012; Valenti et al., 2020; Yao et al., 2021).

One factor likely contributing to this heterogeneity is chronological age. Age-related effects in the neurodevelopmental trajectory of ASD are well documented. Broadly, these include early brain overgrowth, abnormal structural development including morphological anomalies and white matter aberration, and altered neural connectivity (Girault & Piven, 2020). The dMRI literature suggests an atypical developmental trajectory of WM in ASD, whereby early WM abnormalities appear to ameliorate with age. More specifically, group differences in classic diffusion tensor imaging (DTI) metrics (FA, AD, MD, and RD) often “normalize” to a level where these indices of WM microstructure become comparable to a non-clinical cohort during adulthood. Indeed, our own research using similar, tensor-based, protocols in adults with ASD found no differences in WM microstructure compared to a matched NT comparison group (Kirkovski, Enticott, Maller, Rossell, & Fitzgerald, 2015). Moreover, recent work suggests that the pattern, or direction, of microstructural aberration might actually “switch” in ASD (Karahanoğlu et al., 2018). Here, the authors report higher FA and AD in younger (aged 8-15) individuals with ASD compared to a matched NT comparison group, while the opposite pattern was observed for the older (16-25 years) cohort (Karahanoğlu et al., 2018). Longitudinal dMRI research further supports these observations (Andrews et al., 2021; McLaughlin et al., 2018; Travers et al., 2015). Thus, while the developmental profile of WM aberration in ASD is yet to be fully elucidated, there is evidence that this effect alters as a function of age.

Focusing on the CC in particular, similar age-related patterns and differences have been reported in those with ASD. Travers et al. (2015) conducted a longitudinal dMRI investigation of CC microstructure, following up 100 males with ASD and 56 male NT controls, aged between 3 – 40 years, at four time points. The authors report several age-by-group interactions across DTI metrics and CC sub-regions. These effects can be described by decreased FA across the entirety of the CC, divided into the genu, body, and splenium, and increased MD at the splenium in individuals <10 years old. In support of the view that microstructural abnormalities in ASD may dissipate with age, no significant age-related effects were observed in the older (10-20 years, and 20+ years) cohorts (Travers et al., 2015). Alternatively, more recent work from Andrews et al. (2021) supports the ‘switching’ hypothesis of microstructural differences in those with ASD relative to controls. This study followed 125 children with ASD and 69 NT controls aged between 2.5 – 7 years old over 2-3 time points. The authors report that the developmental trajectory of FA at the body and splenium was slower in the ASD cohort, indicating increased FA in younger children with ASD and decreased FA in older children with ASD compared to NT children. At the genu, the developmental trajectory of FA was relatively similar to that of the NT group (Andrews et al., 2021). Given the variability in ages investigated, and that the entire age range investigated by Andrews et al. (2021) fits within the “younger” group of the earlier longitudinal study by Travers et al. (2015), the age at which these “switches” in, or “normalization” of, microstructural development occurs is therefore unclear. It is also important to note that microstructural abnormalities are indeed also reported among some studies investigating adults with ASD (Travers et al., 2012), demonstrating that age is likely one of many possible factors contributing to the observed heterogeneity in WM microstructure in ASD.

While age is a major factor contributing to the heterogenous neuropathophysiology of ASD, some of the heterogeneity observed might be attributable to the previously described DTI-based metrics themselves. These older methods of investigating WM microstructure are limited by their inability to account for multiple-, or crossing-, fibers within a voxel (Farquharson et al., 2013; Jeurissen, Leemans, Tournier, Jones, & Sijbers, 2013). This is an important consideration, particularly for the CC, given the multitude of fibers crossing through this structure due to its role as the primary pathway for interhemispheric signal transfer throughout the brain (De Lacoste et al., 1985; van der Knaap & van der Ham, 2011). Alternatively, a fixel-based analysis (FBA) approach allows for individual fiber populations (called “fixels”) to be investigated *within* each single voxel, thereby properly accounting for crossing-fibers (Dhollander et al., 2021; Raffelt et al., 2015). This approach improves the biological specificity through which WM can be assessed via indices such as fiber density (FD; a microstructural measure of intra-axonal volume), fiber cross-section (FC; an index of fiber bundle morphology), and combined fiber density and cross-section (FDC; a combined metric assessing the influence of both micro- and macrostructural changes in WM) (Raffelt et al., 2015). Only two studies have applied an FBA approach to ASD data, both implicating the CC in their findings of WM neuropathophysiology, but neither of which investigated age-related effects (Dimond et al., 2019; Kirkovski et al., 2020). Among a sample of 14-20 year olds (Dimond et al., 2019) reported reduced FD in the splenium and genu of the CC in individuals with ASD (n=25) compared to NT controls (n=27). Beyond the CC, this study also identified reduced FD at the inferior fronto-occipital fasciculus bilaterally, and the right arcuate and uncinate fasciculi (Dimond et al., 2019). In an older sample aged 19-56 years, our own research identified a non-significant trend toward reduced FD at the posterior midbody and isthmus of the CC among the ASD group (n=25) compared to matched controls (n=24) (Kirkovski et al., 2020).

Leveraging data accessed from the Autism Brain Imaging Data Exchange-II (ABIDE-II) database, here we apply FBA to investigate age-related micro- and macrostructural neuropathophysiology in ASD. The present study, to our knowledge, is the first to apply the FBA framework in this regard. It was hypothesized that greater WM micro- and macrostructural aberration would be present in younger ASD groups, compared to their matched NT counterparts, and relative to older cohorts.

## Materials and Methods

dMRI and anatomical (T1-weighted) image data were obtained from the ABIDE-II (http://fcon_1000.projects.nitrc.org/indi/abide/abide_II.html) database. Five dMRI datasets are available via the ABIDE-II database. Data from one site, New York University Langone Medical Center: Sample 2, did not include a NT comparison group and was not included in this study. In addition, due to missing gradient encoding information, dMRI data from the Institut Pasteur and Robert Debré Hospital site were also not analyzed here. Therefore, dMRI data from 3 sites were analyzed in the present study. These were: New York University Langone Medical Center, Sample 1 (NYU Sample 1), San Diego State University (SDSU), and the Trinity Centre for Health Science (TCD). Children were acclimatized to the scanning environment at each site by undergoing a mock MRI session before the live scan. Given technical challenges, limitations and confounds of analyzing multi-site dMRI data using different acquisition parameters (Zhu et al., 2011), data from each site were analyzed separately. All image processing and analyses were performed via the multi-modal Australian ScienceS Imaging and Visualization Environment (MASSIVE) supercomputing infrastructure (Goscinski et al., 2014).

Due to technical and data quality issues described below, several participants were excluded from analysis. For data included for analysis, participant demographics are detailed, by site, in Table 1.

**Table 1.**
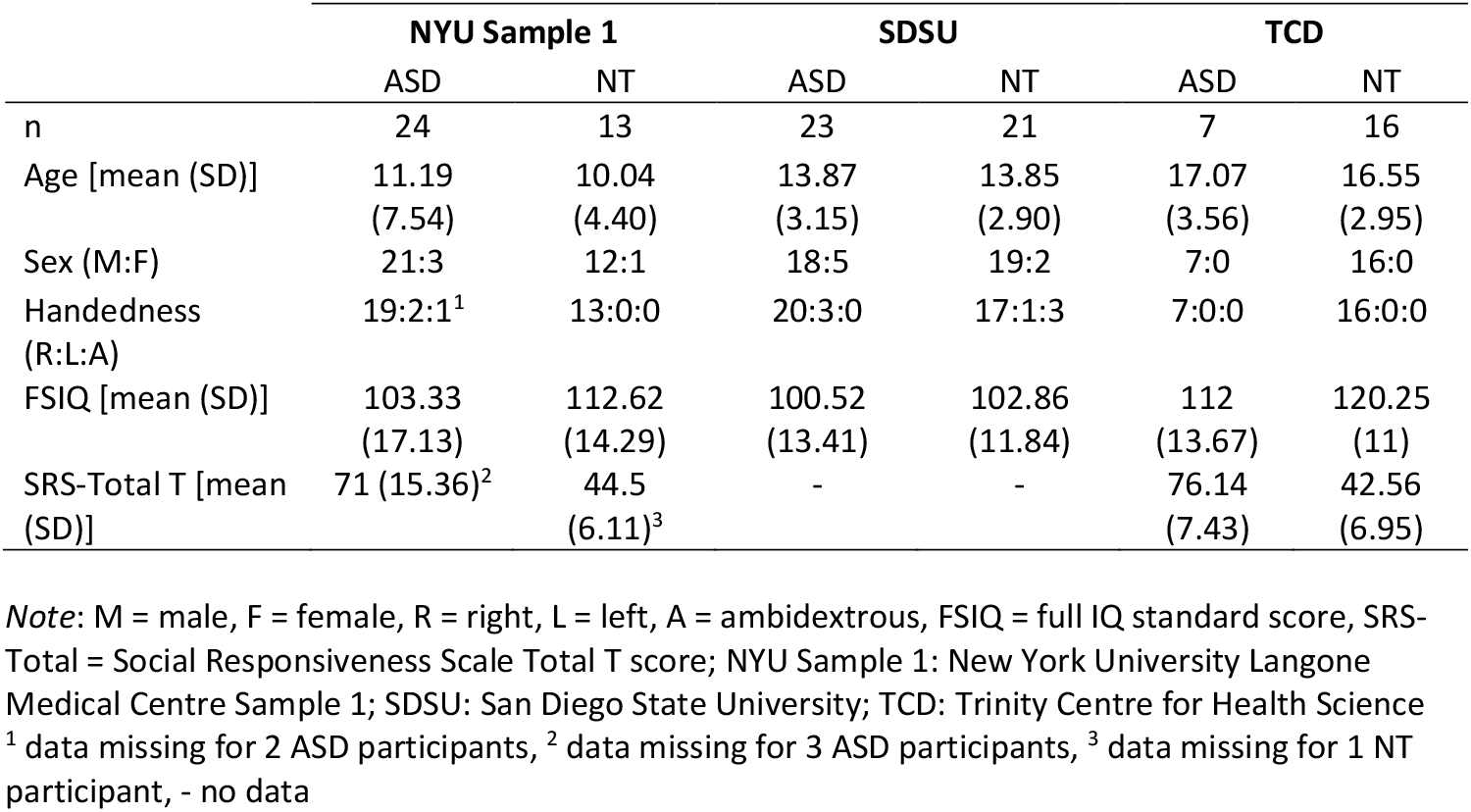
Summary of demographics and participant characteristics by site.

|  | NYU Sample 1 |  | SDSU |  | TCD |  |
| --- | --- | --- | --- | --- | --- | --- |
|  | ASD | NT | ASD | NT | ASD | NT |
| n | 24 | 13 | 23 | 21 | 7 | 16 |
| Age [mean (SD)] | 11.19<br>(7.54) | 10.04<br>(4.40) | 13.87<br>(3.15) | 13.85<br>(2.90) | 17.07<br>(3.56) | 16.55<br>(2.95) |
| Sex (M:F) | 21:3 | 12:1 | 18:5 | 19:2 | 7:0 | 16:0 |
| Handedness<br>(R:L:A) | 19:2:1<sup>1</sup> | 13:0:0 | 20:3:0 | 17:1:3 | 7:0:0 | 16:0:0 |
| FSIQ [mean (SD)] | 103.33<br>(17.13) | 112.62<br>(14.29) | 100.52<br>(13.41) | 102.86<br>(11.84) | 112<br>(13.67) | 120.25<br>(11) |
| SRS-Total T [mean<br>(SD)] | 71 (15.36)<sup>2</sup> | 44.5<br>(6.11)<sup>3</sup> | - | - | 76.14<br>(7.43) | 42.56<br>(6.95) |

*Note:* M = male, F = female, R = right, L = left, A = ambidextrous, FSIQ = full IQ standard score, SRS-Total = Social Responsiveness Scale Total T score; NYU Sample 1: New York University Langone Medical Centre Sample 1; SDSU: San Diego State University; TCD: Trinity Centre for Health Science <sup>1</sup> data missing for 2 ASD participants, <sup>2</sup> data missing for 3 ASD participants, <sup>3</sup> data missing for 1 NT participant, - no data

An overview of the acquisition parameters, as provided to ABIDE-II, used in each scanning site is presented in Table 2. Further site-specific details can be sourced from: http://fcon_1000.projects.nitrc.org/indi/abide/abide_II.html

**Table 2.** Acquisition parameters per site: dMRI and T1.

|  | NYU Sample 1 |  | SDSU |  | TCD |  |
| --- | --- | --- | --- | --- | --- | --- |
| Manufacturer | Siemens Magnetom |  | GE M7R50 |  | Philips Achieva |  |
| Tesla strength | 3 |  | 3 |  | 3 |  |
|  | dMRI | T1 | dMRI | T1 | dMRI | T1 |
| Sequence | EPI | MPRAGE | EPI | FSPGR | EPI | MPRAGE |
| Voxel size (mm3) | 3 x 3 x 3 x 5.2 | 1.3 x 1 x 1.3 | 1.875 x 1.875 x 2 x 1 | - | 1.938 x 1.938 x 2 x 1 | .89 x .89 x .89 |
| Slice thickness (mm) | 3 | 1.33 | 2 | 1 | 2 |  |
| Orientation | Axial | Sagittal | Axial | Sagittal | Axial | Transverse |
| Flip angle | 60 | 7 | - | 8 | 90 | 8 |
| b value (s/mm3) | 1000 | - | 1000 | - | 1500 | - |
| dirs/b=0 | 64/1 | - | 61/1 | - | 61/1 | - |
| Acquisition time (min: sec) | 5:43 | 8:07 | - | - | 24:21.7 | 05:43.6 |
| TE (ms) | 78 | 3.25 | Minimum | Min Full | 79 | shortest |
| TR (ms) | 5200 | 2530 | 8500 | - | 20244 | 8.4 |
| FoV (mm) | 192 x 192 | 256 x 256 | 128 x 128 | 256 x 256 | 248 x 248 | 256 x 256 |

*Note:* NYU Sample 1: New York University Langone Medical Centre Sample 1; SDSU: San Diego State University; TCD: Trinity Centre for Health Science; TE: Echo time; TR: Repetition time; FoV: Field of view; EPI: Echo Planar Imaging; - no data; MPRAGE: Magnetization-prepared 180 degrees radio-frequency pulses and rapid gradient-echo; FSPGR: Fast spoiled gradient-echo resonance.

### dMRI data preparation

Quality assessment was performed by two raters (MK and MS). All steps were performed in MRtrix3Tissue (v5.2.8. https://3tissue.github.io/), a fork of the MRtrix3 software package (Tournier et al., 2019). Prior to processing, the raw data from each scanning site were assessed to ensure they met basic quality checks. Subjects were removed if they were missing dMRI data (NYU: n = 21; SDSU: n = 1; TCD: n = 2) or gradient encoding information (NYU: n = 2). The data for all remaining subjects were then converted from NIfTI to MRtrix3-native format.

### Quality Assessment

Gradient encoding information for all converted images was checked to ensure that the correct orientations were preserved during the conversion, which led to the removal of one subject where this information was corrupted (SDSU: n =1). The data were then visually inspected by both raters for motion artifacts in areas where we would expect the CC to traverse (indicated by the presence of Venetian blinding and/or signal dropout). This resulted in the removal of 48 subjects (NYU: n = 18; SDSU: n = 13; TCD: n = 17). Based on the QC, the final sample consisted of 37 subjects for the NYU (ASD = 24; NT = 13), 44 for the SDSU (ASD = 23; NT = 21), and 23 for the TCD (ASD = 7; NT = 16) scanning sites, respectively.

### Head Motion and Intracranial volume

We derived quantitative estimates of in-scanner head motion and total brain volume for all subjects in our FBA analysis. In-scanner head motion was operationalized as mean frame-wise displacement (FWD) and calculated across each subject’s raw dMRI dataset (excluding the b=0 volume) in FSL (Jenkinson, Beckmann, Behrens, Woolrich, & Smith, 2012) using the approach described by Power, Barnes, Snyder, Schlaggar, and Petersen (2012). Brain volume measures were calculated separately for gray matter (GM), WM and cerebrospinal fluid (CSF) using T1-weighed structural images in FreeSurfer (Fischl, 2012). We also calculated the estimated total intracranial volume (eTIV). Mann-Whitney tests revealed no statistically significant group differences in FWD and brain volume across all scanning sites (see Supplementary Table 1).

### Preprocessing and Fixel-based analysis

Data were preprocessed according to a state-of-the-art workflow for FBA (Dhollander et al., 2021). We applied the same steps across all subjects for each respective scanning site. Briefly, each dataset underwent denoising (Veraart, Fieremans, & Novikov, 2016) and Gibbs unringing (Kellner, Dhital, Kiselev, & Reisert, 2016) before correcting for motion and eddy-current distortions (Andersson & Sotiropoulos, 2016). Tissue-specific response functions for WM, GM and CSF dMRI signals were estimated individually from each subject’s preprocessed image using an unsupervised method (Dhollander, Mito, Raffelt, & Connelly, 2019). These response functions were then averaged across subjects in each cohort to generate group-level response functions (Dhollander et al., 2021).

Data were then upsampled to a voxel size of 1.5 × 1.5 × 1.5 mm^3^ to improve spatial anatomical contrast and single-shell 3-tissue constrained spherical deconvolution (SS3T-CSD) was conducted to obtain WM Fiber Orientation Directions (FODs) for each subject (Dhollander & Connelly, 2016). We further applied group-level intensity normalization to ensure that FOD amplitudes were directly comparable across subjects within each separate (scanning site) dataset. To ensure the biological plausibility of FODs underlying the CC, we then performed an additional quality control step, where FOD maps for each subject were visually inspected to determine whether they had adequately accounted for crossing fibers (particularly within the distal regions of the CC), and their spatial alignment with the direction of diffusion along the CC. All subjects showed plausible FODs and were therefore included in all analyses.

Subjects in each cohort were then registered to an unbiased population template space using non-rigid registration. Subject details for each template image are as follows: (1) NYU = 37 (ASD = 24; NT = 13), (2) SDSU = 44 (ASD = 23; NT = 21), and (3) TCD =23 (ASD = 7; NT = 16). All population templates were further warped to Montreal Neurological Institute (MNI) space in FSL (v6.0.1) to compare regions of significance with those of previous work (Dimond et al., 2019; Kirkovski et al., 2020). Further to this, whole-brain fixel analysis masks were estimated for each template by thresholding peak FOD values to 0.1 for all datasets. Lastly, fixel-wise metrics of FD, FC and FDC were computed across all template fixels for subsequent inferential analysis (Raffelt et al., 2017). FC estimates specifically were log-transformed (logFC) before statistical analysis.

### Tractography

We employed a tract-of-interest approach to evaluate WM micro- and macrostructural differences in the CC across individuals with ASD and NT. The CC was delineated in each population template using TractSeg (Wasserthal, Neher, & Maier-Hein, 2018), an automated method that provides segmentation of 72 major WM tracts learned from a reference dataset of 105 subjects acquired from the Human Connectome Project (Van Essen et al., 2013).

### Connectivity-based fixel enhancement

Group differences in FD, FC and FDC for all tracts-of-interest were conducted using threshold-free Connectivity-based Fixel Enhancement (CFE) (Raffelt et al., 2015). We employed the following default parameters: smoothing = 10 mm full width at half maximum, C = 0.5, E = 2, H = 3. For analyses involving logFC and FDC, we further adjusted for estimated total intracranial volume (eTIV) to account for individual differences in head size on our results. Age was also included as a covariate of interest for all analyses, given its established role in the neuropathophysiology of ASD. All analyses were corrected for multiple comparisons using familywise error rates (FWE). Where significant results were identified (*p*^*FWE*^ < 0.05), tractograms were cropped so that only streamlines traversing through significant fixels were displayed to better ascertain the WM pathways implicated in the neuropathophysiology of ASD.

### Correlations between age and fixel-wise metrics

Based on the analyses described above, where relevant (i.e. where significant differences in fixel-wise metrics were observed between groups) mean FD, logFC or FDC values were extracted (averaged across the CC) and plotted against age for each subject using RStudio v.4.0.5. Due to the relatively small sample sizes within each data set, Spearman’s Rank Order Correlations were used to assess the relationship between age and each of the FBA metrics implicated in the neuropathophysiology of ASD. A p-value of <0.01 was considered statistically significant to account for multiple comparisons, based on a simple Bonferroni correction.

### Code and Data availability

All analysis scripts used in the present study are publicly available via: https://github.com/MervSingh/ABIDE-II_FBA.git. The ABIDE-II datasets are open source and can be downloaded from the following weblink: https://fcon_1000.projects.nitrc.org/indi/abide/.

## Results

### Comparison of white matter micro- and macrostructure between groups

#### Younger adolescents (NYU site)

FBA revealed significantly decreased logFC and FDC among the ASD group compared to NT controls, particularly around the midbody, isthmus and splenium regions of the CC. A non-significant fixel cluster (*p*^*FWE*^ = <.08) indicative of reduced FD among the ASD group was also identified at these regions. Within this cluster, only one fixel remained statistically significant at (*p*^*FWE*^ = <.05). Streamline segments of implicated fixels are presented in Figure 1. No regions with increased FD, FC, or FDC among the ASD group were identified.

**Figure 1.**
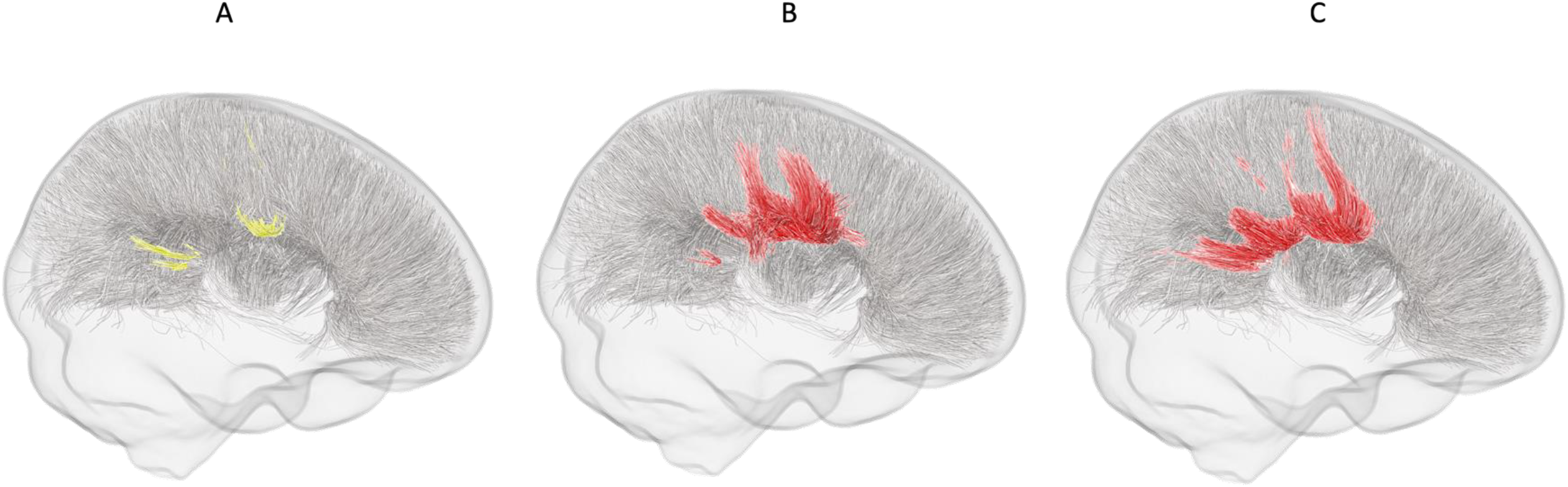
Younger adolescents (NYU site). Graphical representation of streamlines for fixels showing a trend-level reduction in (A) FD, and statistically significant (*p*^*FWE*^ <.05) reductions in (B) logFC, and (C) FDC in the ASD group compared to NT controls. Trend level streamlines are represented in yellow, while significant streamlines are red.

#### Older adolescents (SDSU site)

FBA revealed a statistically significant reduction in FD in the ASD group in a small region of the splenium. A significant reduction in FDC at the midbody was also identified in the ASD group compared to controls. Streamline segments of implicated fixels are presented in Figure 2. No regions with increased FD, FC, or FDC among the ASD group were identified.

**Figure 2.**
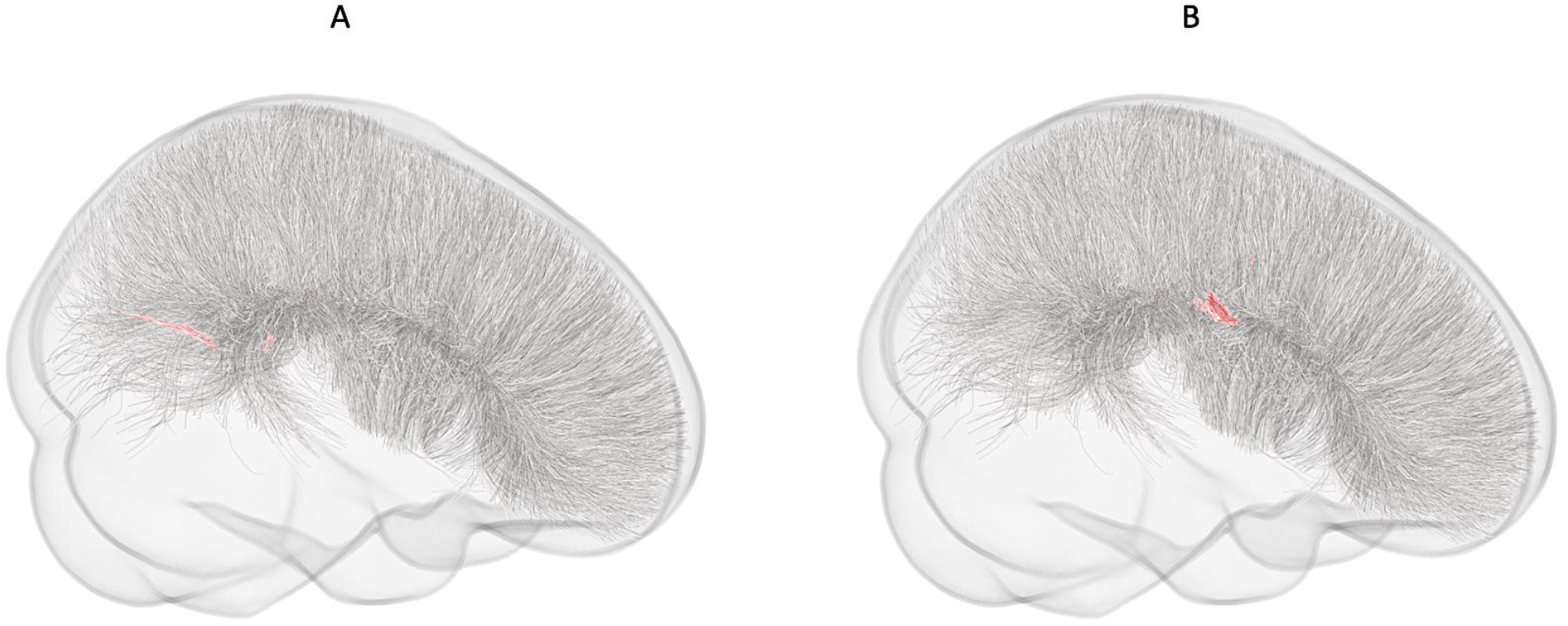
Older adolescents (SDSU site). Graphical representation of streamlines for fixels showing statistically significant (*p*^*FWE*^ <.05) reductions in (A) FD, and (B) FDC in the ASD group compared to NT controls.

#### Older adolescents/young adults (TCD site)

FBA analysis did not reveal any significant differences in WM micro- or macro-structure between ASD participants and NT controls within this data set. A non-significant trend (*p*^*FWE*^ = 0.08), however, indicative of reduced FD around the isthmus/splenium in the ASD group compared to NT controls was observed (Figure 3). Within this cluster, only a single fixel remained statistically significant at (*p*^*FWE*^ = <.05). No regions with increased FD, FC, or FDC among the ASD group were identified.

**Figure 3.**
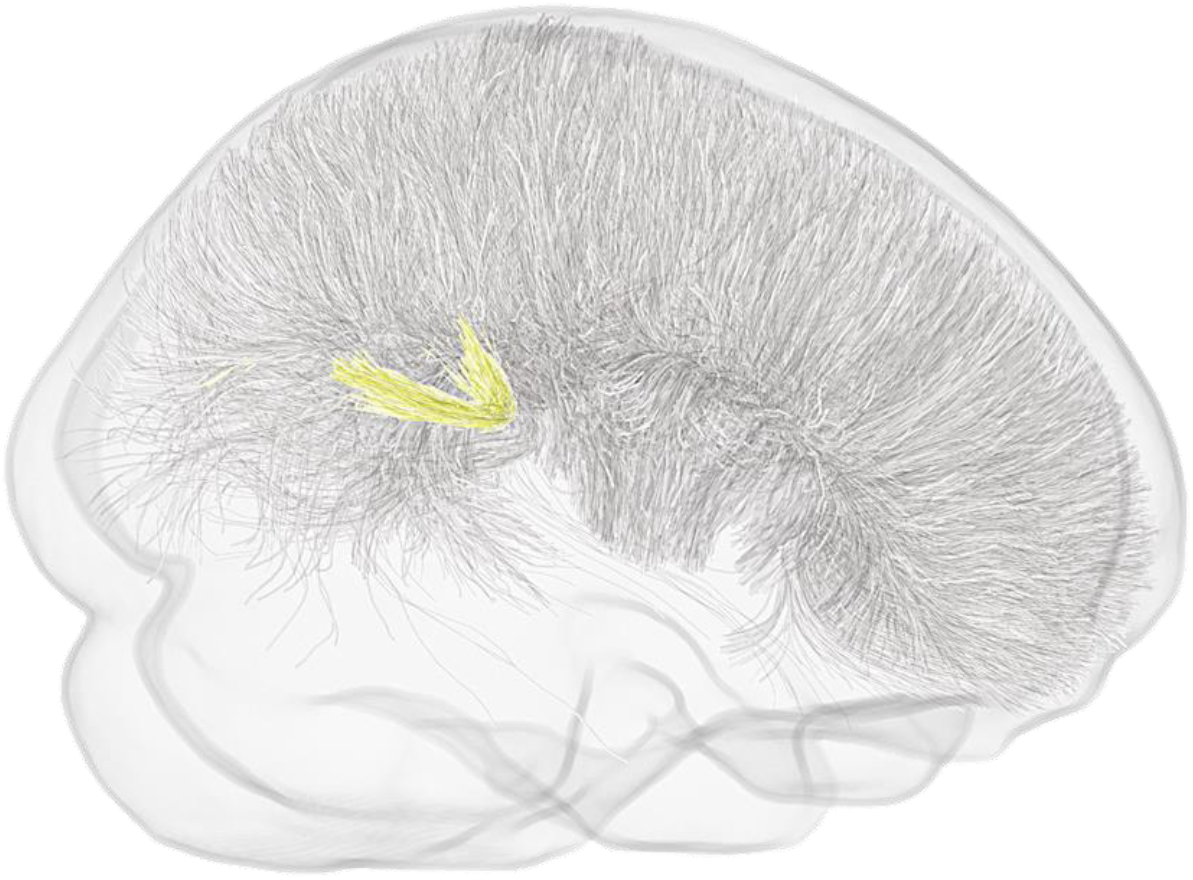
Older adolescents/young adults (TCD site). Graphical representation of streamlines for fixels showing a non-significant trend (p^FWE^ = 0.08) toward reductions in FD among the ASD group compared to NT controls. Trend level streamlines are represented in yellow.

### The relationship between age and white matter micro- and macro-structure

#### Younger adolescents (NYU site)

Spearman’s Rank Order Correlations revealed a moderate positive relationship between age and mean logFC (r = .43, *p* = .009) and mean FDC (r = .52, *p* = .001) for the NYU site. When data were stratified by diagnosis, a similar pattern was observed for the ASD group between age and mean logFC (r = .56, *p* = .005), and also age and mean FDC (r = .54, *p* = .006), refer to Figure 4. There was no relationship between age and either of the relevant fixel-wise metrics for the NT group at this site.

**Figure 4.**
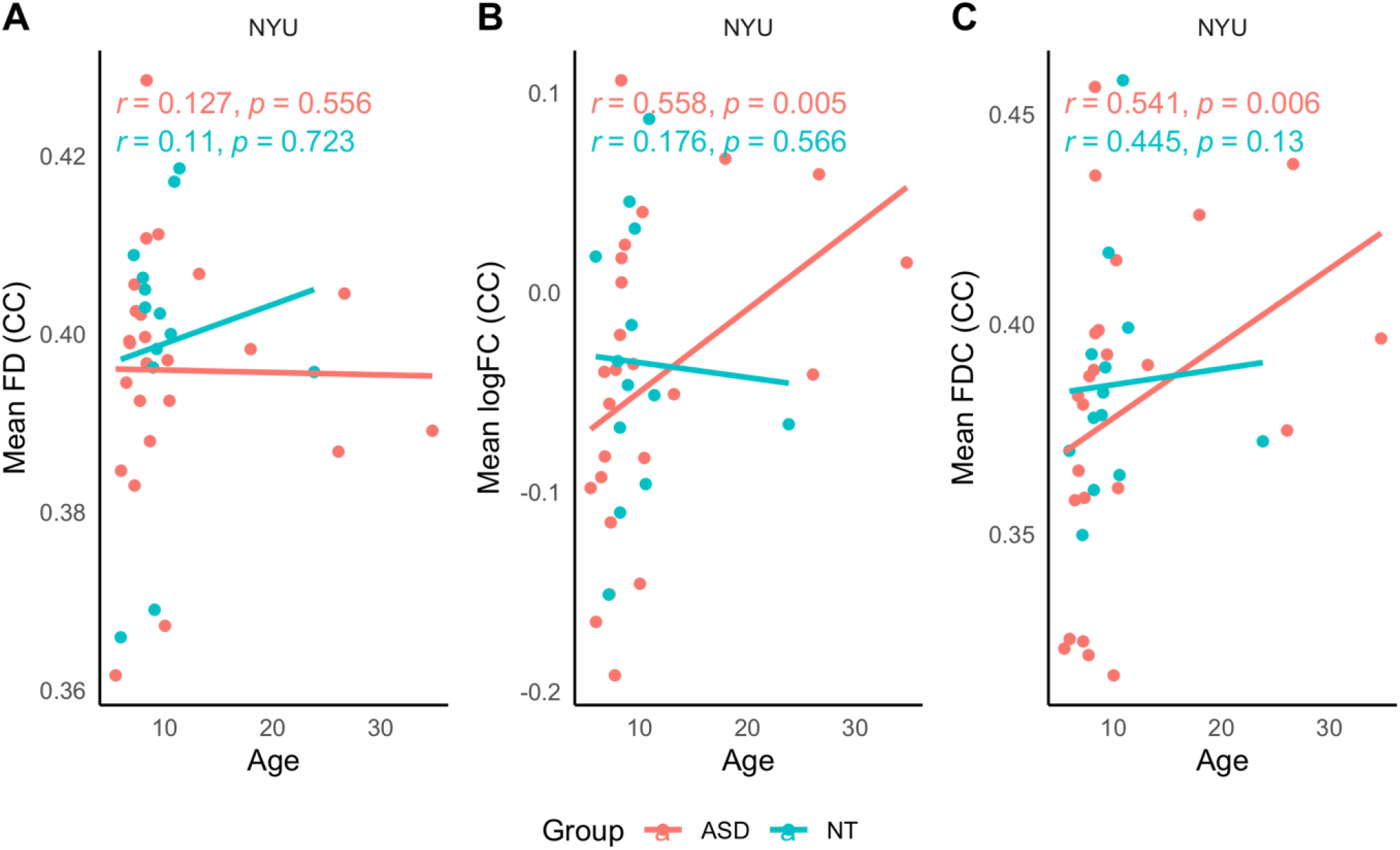
Younger adolescents (NYU site). Scatterplot depicting the relationship between age and mean FD, mean logFC and mean FDC of the CC, stratified by diagnosis.

#### Older adolescents (SDSU site)

Spearman’s Rank Order Correlations revealed no significant relationships between age and mean FD or mean FDC across the whole sample, nor when data were stratified by diagnosis. Scatter-plots are presented in Figure 5.

**Figure 5.**
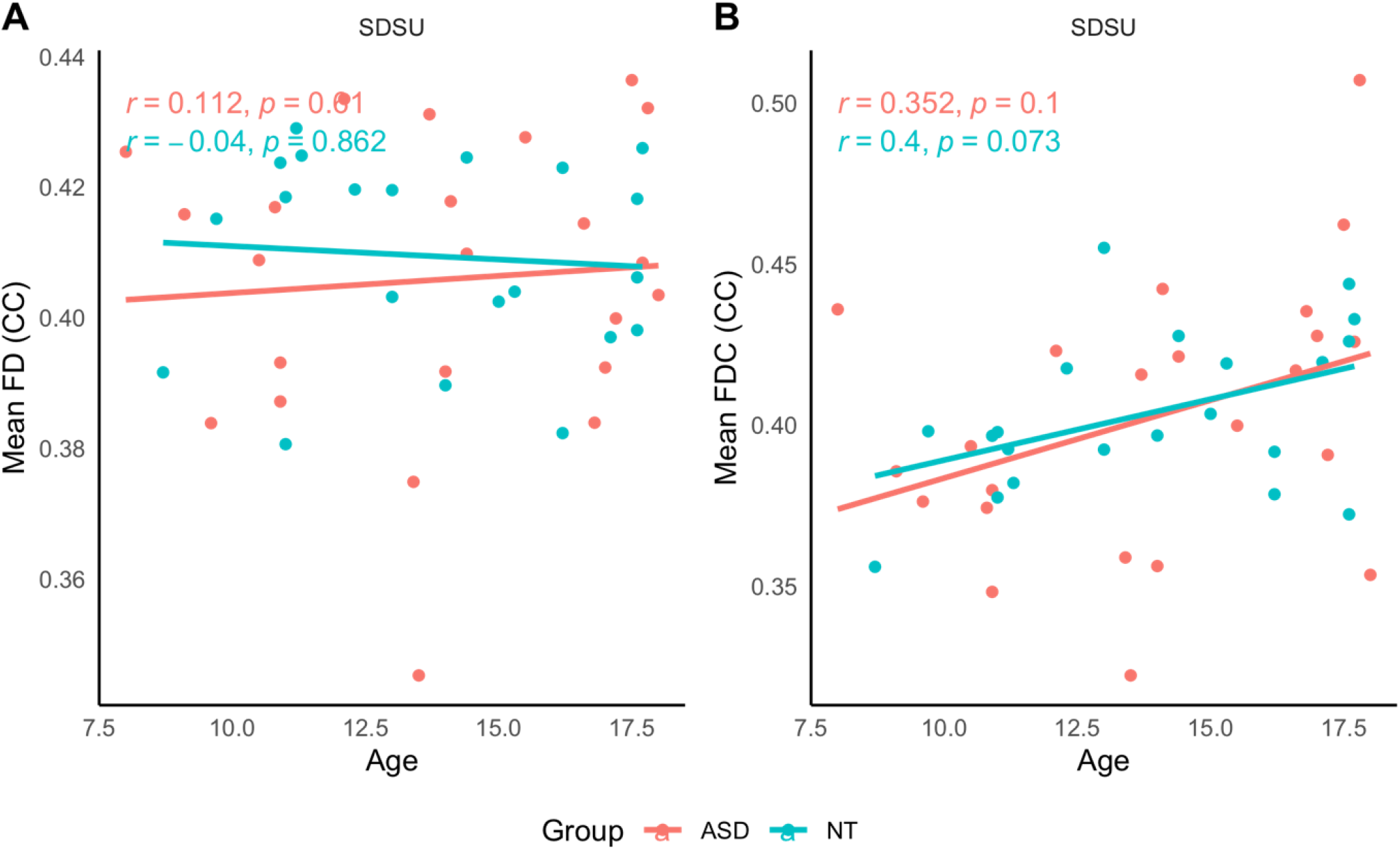
Older adolescents (SDSU site). Scatterplot depicting the relationship between age and mean FD and mean FDC of the CC, stratified by diagnosis.

#### Older adolescents /young adults (TCD site)

Across the entire TCD dataset, there was no statistically significant relationship between age and mean FD. Similarly, there were no relationships between these variables when data were stratified by diagnosis. Refer to Figure 6.

**Figure 6.**
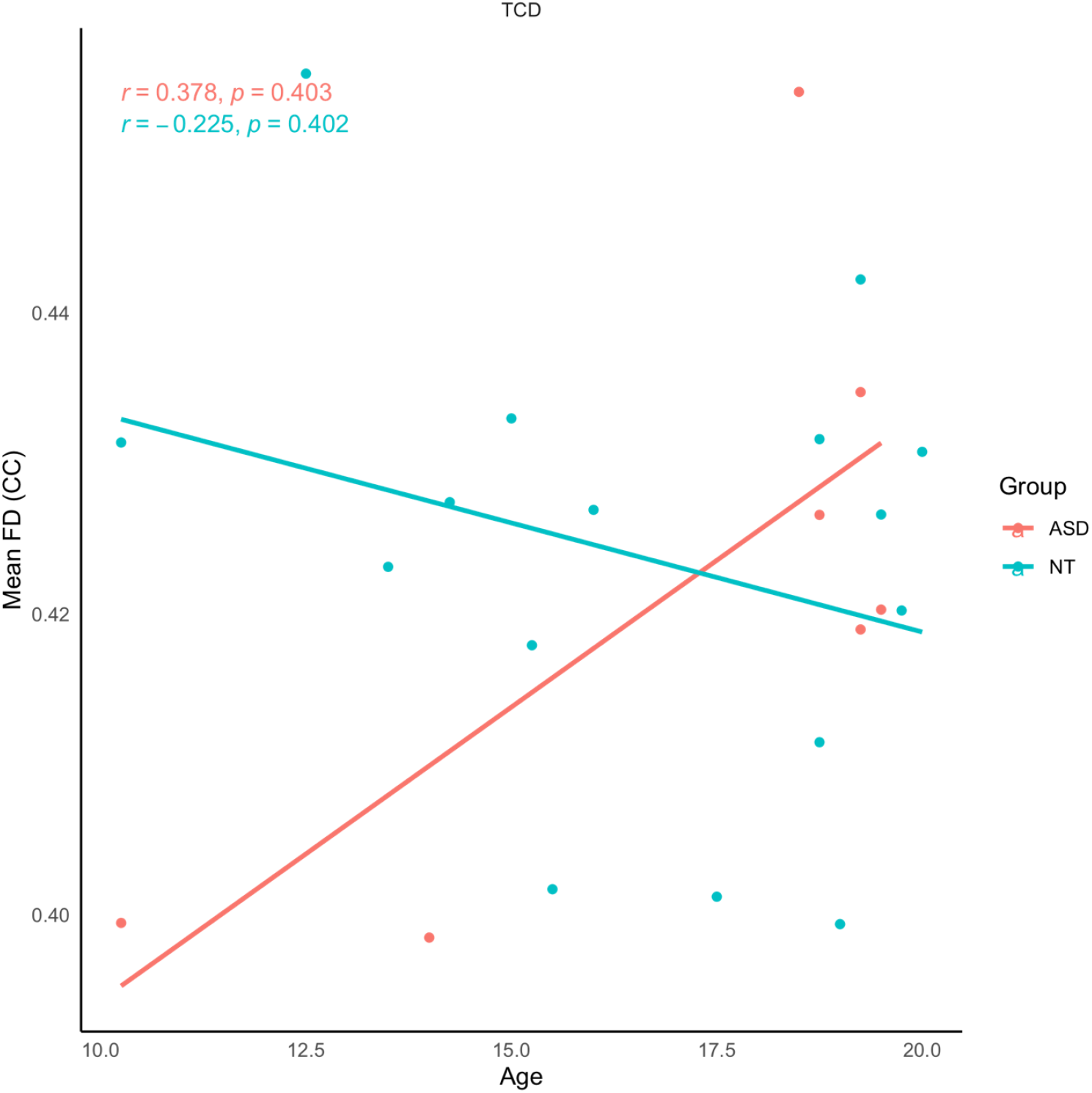
Older adolescents /young adults (TCD site). Scatterplot depicting the relationship between age and mean FD of the CC, stratified by diagnosis.

## Discussion

We applied fixel-based analysis (FBA) to data from three sites obtained from the ABIDE-II database to investigate age-related WM micro- and macrostructural pathology in adolescents and young adults with ASD compared to matched controls. As hypothesized, the youngest ASD cohort (based on the NYU site) showed the greatest WM aberration compared to their NT counterparts. Differences were identified in micro- and macrostructural measures, implicating the midbody, isthmus and splenium. Less, albeit significant, aberration was identified in overlapping regions in the marginally older cohort (SDSU). No significant WM aberration was identified among the oldest cohort (TCD), though there was a non-significant trend towards reduced FD around the isthmus/splenium region in the ASD group. To reconcile the findings across the three sites, we provide evidence of abnormality across WM micro- and macrostructure and affecting a greater proportion of the CC in younger, compared to older adolescent and young adult cohorts. Among the older cohort, aberration was observed in microstructure (FD) only, and was more spatially localized.

Data from the NYU site showed that young adolescents with ASD (average age 11.19 [SD=7.54]) had significantly reduced FC, and FDC compared to matched NT controls (mean age 10.04 [SD = 4.40]). A non-significant trend indicative of reduced FD in the ASD group was also observed in a small fixel cluster. Statistically significant reductions in FD and FDC were, however, identified in overlapping regions in the slightly older ASD group based on SDSU data. As a reminder, reductions in FD might reflect axonal loss or displacement, whereas FC provides a fixel level index of fiber bundle volume relative to the study-specific template (Dhollander et al., 2021; Raffelt et al., 2015; Raffelt et al., 2017). FDC, however, is arguably a more sensitive (and less specific) measure derived from a combination of FD and FC. As explained by Dhollander et al. (2021), there are several drawbacks to interpreting these metrics alone (i.e., each metric in isolation, without context from the other metrics). Therefore, when considering the results presented here, despite non-significant FD outcome, it is reasonable to conclude that both WM micro- and macrostructure is impaired in young adolescents with ASD.

Among the older adolescent/young adult cohort (TCD), there was trend-level evidence of reduced FD at the isthmus/splenium in the ASD group. This finding is in line with our earlier FBA investigation of WM neuropathophysiology in adults with ASD (Kirkovski et al., 2020), where a trend towards reduced FD at the posterior midbody/isthmus was identified among the ASD group. Using diffusion kurtosis imaging (DKI) Sui et al. (2018) provide further evidence aligned with these findings. Specifically, the authors report evidence of reduced axonal density and a reduction in the number of intra-axonal structures at the midbody, isthmus, and splenium of young adults (18-25 years) with ASD compared to matched controls. These findings represent an emerging pattern whereby WM aberration appears concentrated to posterior regions of the CC among older ASD cohorts. In their longitudinal study, Andrews et al. (2021), demonstrate slower development of the splenium of the CC in children with ASD. In considering that several temporal and parietal projections pass through this region of the CC (De Lacoste et al., 1985; Hofer & Frahm, 2006), and the critical role of these cortices in social understanding, a fundamental criterion of ASD (Patriquin, DeRamus, Libero, Laird, & Kana, 2016), further investigation into the clinical relevance of this finding is warranted.

To better understand the role of age in these findings, correlation analyses for the youngest cohort revealed an age-related increase in these metrics among the ASD group only. No significant relationships were identified among any of the older cohorts. This could be interpreted to reflect the neuropathophysiological “normalization” described previously. The concept of “normalization” of neuropathophysiology in ASD comes from early research investigating brain morphology. For over two decades, it has been understood that a pattern of early brain overgrowth in young children with ASD is followed by a size-related “normalization” of these regions in older children (Courchesne, 2004; Courchesne, Campbell, & Solso, 2011; Courchesne & Pierce, 2005). It is important to note, however, that this finding is not universally observed. In a large replication study, Yankowitz et al. (2020) report enlarged brain volume in ASD, from childhood through to adulthood.

Less is understood about the structural and functional overlap in age-related “normalization” of neuropathophysiology in ASD. Using concurrent transcranial magnetic stimulation and electroencephalography (TMS-EEG) Jarczok et al. (2016) report similar maturation effects of interhemispheric signal propagation, indicative of interhemispheric connectivity, in adolescents and young adults with and without ASD. Though these findings might broadly be interpreted to suggest that measures of CC function are not consistent with structural imaging findings, an important consideration here is that interhemispheric signal propagation was measured from the primary motor cortex. We observed some degree of “normalized” WM, primarily around the midbody regions of the CC, fibers of which project into motor, somatosensory and posterior parietal regions (Hofer & Frahm, 2006). This, therefore, might explain the findings reported by Jarczok et al. (2016). While not statistically significant, the present results, and the results of our previous FBA investigation (Kirkovski et al., 2020) indicate that some abnormalities might remain at more posterior regions of the CC (isthmus and splenium), projecting in temporal and parietal structures (Hofer & Frahm, 2006), as mentioned above. Indeed, while not investigating the effects of age, Kana, Libero, Hu, Deshpande, and Colburn (2014) report reduced temporo-parietal FA and blood oxygen level dependent (BOLD) response in ASD during a task measuring social understanding. Taken together, these findings suggest that the structural/functional overlap is region specific.

Vital to consider is the clinical impact of this neuropathophysiological “normalization” on related behavioral symptomatology, but research that has systematically and directly investigated this is scarce. Clinically, or behaviorally, there is evidence to suggest that the severity of ASD symptomatology might abate slightly with age, particularly for those with a less severe clinical profile (Seltzer et al., 2003; Seltzer, Shattuck, Abbeduto, & Greenberg, 2004; Taylor & Seltzer, 2010).

### Strengths, limitations and future directions

A key strength of the present study is that FBA, a cutting-edge framework for dMRI analysis, capable of accounting for crossing fibers, has been consistently applied with identical processes, to investigate the role of age on WM pathophysiology in ASD across data from three separate sites. There are, however, important limitations to be acknowledged. Firstly, as described above, combining dMRI data across different acquisition protocols and/or sites to drive more statistically powerful analyses is not straightforward. Even with efforts to harmonize data, this risks introducing biases in the analysis. It might not even be reasonable or sensible to attempt combining dMRI data acquired at, for example, different b-values (diffusion weightings), as such data would be sensitized to fundamentally different aspects of microstructure. Further to this point, the samples available from each site in our dataset are independently small, particularly for demanding neuroimaging analyses such as FBA. This is a critical consideration, particularly for the TCD sample. Hence, while cautious in our interpretations regarding the non-significant outcomes identified based on these data, they nevertheless support the notion that ASD neuropathophysiology might reduce with age. Another related consideration is that while these datasets have been used to delineate different age groups, separate comparisons limit our ability to account for characteristics inherent to each sample or study design. Finally, regarding the data, we acknowledge the overlap in age across our samples, particularly that the age range of our youngest (average age) sample (NYU) did overlap with the two older sites.

An important methodological consideration is that these data are not entirely optimized for FBA analysis, but rather had likely been acquire with DTI-based modeling approaches in mind. Higher b-values (e.g., 3000 s/mm^2^) during acquisition yield a better contrast to noise ratio, which promotes better FOD quality from which the FD metric is derived. The FD metric itself is also more specific to intra-axonal volume (which it is meant to represent) at such higher b-values. Though still appropriate for FBA analysis in principle, given the lower b-values of the data presented here we took additional steps to ensure FOD quality (Dhollander et al., 2021), as described in the methods section. Nevertheless, the available data provides important insights into the age-related micro-and macrostructural neuropathophysiology of ASD, setting up the basis for large scale and technically optimized investigation.

Finally, we were limited to the available data obtained from the ABIDE-II database. Several factors other than age are known or exceedingly likely to contribute to the heterogeneity observed in ASD. Our own research (Kirkovski, Enticott, & Fitzgerald, 2013; Kirkovski, Enticott, Hughes, Rossell, & Fitzgerald, 2016; Kirkovski et al., 2015; Kirkovski et al., 2020; Kirkovski, Suo, Enticott, Yücel, & Fitzgerald, 2018), for example, highlights biological sex as a factor linked to the neuropathophysiology of ASD. The present data, however, did not allow for this to be explored. In addition, the roles of clinical features (Giuliano et al., 2018) and symptom severity (Andrews et al., 2021) should also be systematically evaluated by future research.

## Summary and conclusion

In summary, the CC is one of the most commonly implicated WM tracts in research investigating the neuropathophysiology of ASD. Though there are several reports of age-related “normalization” (that is, greater neurobiological abnormality among younger ASD cohorts) using traditional dMRI measures, here we present the first study to use FBA to investigate this phenomenon. We provide evidence that WM aberration is greatest in younger ASD cohorts, and the extent of this aberration is reduced, or altered, in older cohorts. Further investigation of WM pathology in a broader age-range, as well as in larger cohorts, using this method will be of great benefit to the field. Future research should also consider other factors known to contribute to the heterogeneity of ASD, such as biological sex, symptomatology, and cognitive ability.

## Supporting information

(see Supplementary Table 1)

## Acknowledgements

We thank the participants who took part in the research and sites who made the data available for use via the ABIDE-II database.

## Funding and disclosures

This research was funded by an Alfred Deakin Post-Doctoral Research Fellowship (Deakin University) held by MK. MS is supported by the Deakin University Postgraduate Research Scholarship (DUPR) PGE was supported by a Future Fellowship from the Australian Research Council (FT160100077)

The researchers declare no conflict of interest.

## Notes

### Competing Interest Statement

The authors have declared no competing interest.

### Author Declarations

Only data that were openly available to the public before the initiation of the study were used. Data were obtained from the ABIDE-II (http://fcon_1000.projects.nitrc.org/indi/abide/abide_II.html) database.

