## Supplementary material for "An investigation of age-related neuropathophysiology in autism spectrum disorder using fixel-based analysis of corpus callosum white matter micro- and macrostructure": (see Supplementary Table 1)

Supplementary Table 1. Descriptive statistics and group comparisons for head motion and brain volume metrics by site

| | ASD | | | | | | | NT | | | | | | | | |
|---|---|---|---|---|---|---|---|---|---|---|---|---|---|---|---|---|
| | n | M | SD | Median | Min | Max | Mean Rank | n | M | SD | Median | Min | Max | Mean Rank | *U* | *p* |
| **NYU** | 24 | | | | | | | 13 | | | | | | | | |
| FWD | | 0.478 | 0.087 | 0.480 | 0.325 | 0.678 | 18.92 | | 0.479 | 0.075 | 0.493 | 0.353 | 0.601 | 19.15 | 158.00 | 0.96 |
| T-GM | | 754833.74 | 64896.41 | 768187.81 | 626678.07 | 837179.28 | 18.38 | | 758833.93 | 70347.38 | 759934.10 | 626678.07 | 837179.28 | 20.15 | 141.00 | 0.65 |
| T-WM | | 417032.57 | 50376.29 | 416418.30 | 342719.50 | 509573.74 | 18.04 | | 426699.31 | 43298.98 | 427810.07 | 342719.50 | 509573.74 | 20.77 | 133.00 | 0.48 |
| CSF | | 911.71 | 195.92 | 866.00 | 723.70 | 1310.90 | 17.33 | | 996.17 | 205.43 | 927.10 | 723.70 | 1310.90 | 22.08 | 116.00 | 0.21 |
| eTICV | | 1453689.36 | 135524.90 | 1450597.47 | 1301968.95 | 1715666.05 | 18.29 | | 1494756.52 | 132596.59 | 1467770.56 | 1301968.95 | 1715666.05 | 20.31 | 139.00 | 0.60 |
| **SDSU** | 23 | | | | | | | 21 | | | | | | | | |
| FWD | | 0.433 | 0.068 | 0.432 | 0.267 | 0.547 | 23.09 | | 0.439 | 0.069 | 0.422 | 0.345 | 0.611 | 21.86 | 228.00 | 0.75 |
| T-GM | | 791323.91 | 55322.80 | 800835.03 | 674370.20 | 877581.46 | 23.83 | | 782327.14 | 53539.69 | 779748.78 | 710189.60 | 884525.83 | 21.05 | 211.00 | 0.48 |
| T-WM | | 440507.12 | 55748.39 | 435542.93 | 367041.76 | 549514.18 | 21.78 | | 442515.42 | 45567.59 | 441287.49 | 358120.55 | 520808.52 | 23.29 | 225.00 | 0.71 |
| CSF | | 925.61 | 210.13 | 968.80 | 515.80 | 1324.60 | 22.78 | | 912.58 | 178.41 | 932.30 | 546.00 | 1287.40 | 22.19 | 235.00 | 0.89 |
| eTICV | | 1549174.80 | 170876.11 | 1541564.40 | 1232284.58 | 1941915.38 | 21.48 | | 1569587.65 | 152385.10 | 1562880.07 | 1329507.79 | 1871789.47 | 23.62 | 218.00 | 0.59 |
| **TCD** | 7 | | | | | | | 16 | | | | | | | | |
| FWD | | 0.656 | 0.166 | 0.580 | 0.476 | 0.866 | 13.14 | | 0.624 | 0.119 | 0.578 | 0.461 | 0.849 | 11.50 | 48.00 | 0.62 |
| T-GM | | 759011.86 | 80691.45 | 766470.72 | 632244.55 | 873391.90 | 9.57 | | 788927.48 | 57899.52 | 791076.47 | 657228.52 | 857464.50 | 13.06 | 39.00 | 0.28 |
| T-WM | | 496438.98 | 47300.70 | 485230.28 | 444127.92 | 589941.33 | 10.00 | | 509604.63 | 41934.33 | 502250.42 | 448930.03 | 602206.99 | 12.88 | 42.00 | 0.38 |
| CSF | | 895.47 | 126.61 | 866.70 | 780.00 | 1140.40 | 12.43 | | 909.41 | 216.71 | 844.55 | 616.00 | 1258.30 | 11.81 | 53.00 | 0.87 |
| eTICV | | 1487050.32 | 139790.86 | 1470127.90 | 1310174.24 | 1644078.56 | 8.71 | | 1603578.81 | 103858.69 | 1589736.94 | 1420063.99 | 1813626.32 | 13.44 | 33.00 | 0.14 |

Note. ASD = autism spectrum disorder, NT = neurotypical, NYU = New York University, SDSU = San Diego State University, TCD = Trinity Centre for Health Science, FWD = mean framewise displacement, T-GM = total gray matter (mm^3), T-WM = total white matter (mm^3), CSF = cerebrospinal fulid (mm^3), eTICV = estimated total intracranial volume (mm^3).
